# Evidence of gender bias in the diagnosis and management of COVID-19 patients: A Big Data analysis of Electronic Health Records

**DOI:** 10.1101/2020.07.20.20157735

**Authors:** Julio Ancochea, Jose L. Izquierdo, Savana COVID-19 Research Group, Joan B. Soriano

**Author notes:** **Corresponding author full contact details:** Dr. Joan B Soriano, MD, PhD, FERS, FCCP, Servicio de NeumologÍa, Hospital Universitario de la Princesa, UAM, Diego de León 62, 28005-Madrid, Spain, Cellular. Savana COVID-19 Research Group are: **Ignacio H. Medrano, MD; Alberto Porras, MD, PhD; Marisa Serrano, PhD; Sara Lumbreras, PhD, Universidad Pontificia Comillas (ORCID: 0000-0002-5506-9027); Carlos Del Rio-Bermudez, PhD (ORCID: 0000-0002-1036-1673); Stephanie Marchesseau, PhD; Ignacio Salcedo; Imanol Zubizarreta; Yolanda González, PhD.**.

## Abstract

**Background:** It remains unknown whether the frequency and severity of COVID-19 affect women differently than men. Here, we aim to describe the characteristics of COVID-19 patients at disease onset, with special focus on the diagnosis and management of female patients with COVID-19.

**Methods:** We explored the unstructured free text in the electronic health records (EHRs) within the SESCAM Healthcare Network (Castilla La-Mancha, Spain). The study sample comprised the entire population with available EHRs (1,446,452 patients) from January 1^st^ to May 1^st^, 2020. We extracted patients’ clinical information upon diagnosis, progression, and outcome for all COVID-19 cases.

**Results:** A total of 4,780 patients with a test-confirmed diagnosis of COVID-19 were identified. Of these, 2,443 (51%) were female, who were on average 1.5 years younger than males (61.7±19.4 vs. 63.3±18.3, p=0.0025). There were more female COVID-19 cases in the 15-59 yr.-old interval, with the greatest sex ratio (SR; 95% CI) observed in the 30-39 yr.-old interval (1.69; 1.35-2.11). Upon diagnosis, headache, anosmia, and ageusia were significantly more frequent in females than males. Imaging by chest X-ray or blood tests were performed less frequently in females (65.5% vs. 78.3% and 49.5% vs. 63.7%, respectively), all p<0.001. Regarding hospital resource use, females showed less frequency of hospitalization (44.3% vs. 62.0%) and ICU admission (2.8% vs. 6.3%) than males, all p<0.001.

**Conclusion:** Our results indicate important sex-dependent differences in the diagnosis, clinical manifestation, and treatment of patients with COVID-19. These results warrant further research to identify and close the gender gap in the ongoing pandemic.

## INTRODUCTION

As of July 2020, the World Health Organization (WHO) has declared that the coronavirus disease 2019 (COVID-19) pandemic is far from controlled. The cumulative number of confirmed COVID-19 cases across 216 countries, areas, or territories worldwide amounts to over 11,874,226, and 545,481 confirmed deaths have been reported to date.^1^ Record daily numbers of both infections and casualties are seen in many countries, with many of them already experiencing ‘second waves’ after lockdowns lift.^2^

Ever since COVID-19 was initially identified on December 31, 2019 in Wuhan (Hubei Province, China)^3^, there remain many unknowns regarding the epidemiology, clinical characteristics, prognosis, and management of the disease.^4^ Although substantial efforts have been aimed at improving our clinical understanding of the disease, less is known about the gendered impact of the current pandemic. Indeed, investigating sex- and gender-related issues in healthcare is an ongoing and unmet need,^5^ and it is considered a research priority issue within the WHO’s Sustainable Development Goals, a strategic opportunity to promote human rights, and achieve health for all.^6^

Characterizing the extent to which COVID-19 impacts women and men differently is of vital importance to better understand the consequences of the pandemic and to design equitable health policies and effective therapeutic strategies. In this line, recent evidence suggests that there are indeed sex differences in the clinical outcomes of COVID-19.^7,8,9^ Some hypotheses underscore the influence of hormonal factors,^10^ immune response,^11^ differential distribution of the ACE-2 receptors, and smoking habits,^12^ among others.^13^

To further characterize the gendered impact of COVID-19, here we aimed to address whether the frequency and severity of COVID-19 affect women differently than men. In addition, we sought to explore the factors underlying these differences. To achieve these goals, we used big data analytics and artificial intelligence to explore the unstructured, free-text clinical information captured in the electronic health records (EHRs) of a large series of test-confirmed COVID-19 cases.

## METHODS

This study is part of the BigCOVIData initiative^14^ and was conducted in compliance with legal and regulatory requirements.^15^ This study was classified as a ‘non-post-authorization study’ (EPA) by the Spanish Agency of Medicines and Health Products (AEMPS), and was approved by the Research Ethics Committee at the University Hospital of Guadalajara (Spain). We have followed the STrengthening the Reporting of OBservational studies in Epidemiology (STROBE) guidance for reporting observational research.^16^

### Study design, data source, and patient population

This was a retrospective, multicenter study using secondary free-text data from patients’ EHRs within the SESCAM Healthcare Network in Castilla-La Mancha, Spain. Data was retrieved from all available departments, including inpatient hospital, outpatient hospital, and emergency room, for virtually all types of provided services in each participating hospital. The study period was January 1, 2020 – May 1, 2020.

The study database was fully anonymized and aggregated, so it did not contain patients’ personally identifiable information. Given that clinical information was handled in an aggregate, anonymized, and irreversibly dissociated manner, patient consent regulations do not apply to the present study.

The study sample included all patients in the source population with test-confirmed COVID-19 (mainly PCR+ but also IgG/IgM+).

### Extracting free-text from EHRs: EHRead^®^

To meet the study objectives we used *EHRead*^2^, a technology developed by SAVANA that applies Natural Language Processing (NLP), machine learning, and deep learning to access and analyze the unstructured, free-text information jotted down by health professionals in EHRs. The process used for the extraction of clinical data by *EHRead* has been previously described.^17^ Briefly, all extracted clinical terms are standardized according to a unique terminology. This custom-made terminology is based on SNOMED-CT and includes more than 400,000 medical concepts, acronyms, and laboratory parameters aggregated over the course of five years of free-text mining. These clinical entities are detected in the unstructured free text are then classified based on EHRs’ sections using a combination of regular expression rules and machine learning models. Deep learning (CNN) classification methods, which rely on word embeddings and context information, are also used to determine whether the clinical information is expressed in terms of negative, speculative, or affirmative statements.

### Internal validation

For particular cases where extra specifications are required (e.g., to differentiate COVID cases from other mentions of the term related to fear of the disease or potential contact), the detection output was manually reviewed in more than 5000 reports to avoid any ambiguity associated with free-text reporting. All NLP deep learning models used here were validated using the standard training/validation/testing approach; we used a 75/12/13 split ratio in the available annotated data (between 2,000 and 3,000 records, depending on the model) to ensure efficient generalization on unseen cases. For the linguistic validation of analyzed variables regarding COVID-19 mentions, signs/symptoms (e.g., dyspnea, tachypnea, pneumonia), laboratory values (e.g., ferritin, LDH) and treatments (e.g., hydroxychloroquine, cyclosporine, Lopinavir/Ritonavir) we obtained F-scores (the harmonic mean between precision and recall) greater than 0.80 in all cases. However, the validation of ‘PCR-confirmed COVID-19’ returned a F-score of 0.64; although the precision in the identification of this concept was very high (0.90), the recall value was 0.5. This means that even though our model accurately identifies PCR+ cases (i.e., very low number of false positives), the prevalence data reported here may be underestimated.

### Data Analyses

We generated frequency tables to display the information regarding comorbidities, symptoms, and other categorical variables. Continuous variables (e.g., age) were described using summary tables containing mean, standard deviation, median, minimum and maximum values, and quartiles for each variable. To test for possible statistically significant differences in the distribution of categorical variables between males and females, we used Yates-corrected chi^2^ tests for percentages or analysis of variance for normally distributed continuous variables. Sex ratios and their 95% CIs of several epidemiological and clinical indicators are presented. To determine whether the sex ratios of confirmed COVID-19 cases significantly varied across time, we performed linear regression analyses to test the null hypothesis that the slope is equal to zero. All statistical inferences were performed at the 5% significance level using 2-sided tests or 95% CIs.

## RESULTS

From a source population of 2,045,385 individuals, we extracted and analyzed the clinical information of 1,446,452 patients with available EHRs from January 1^st^ to May 1^st^, 2020. Among these, we then retrieved the clinical information upon diagnosis, progression, and outcome for 4,780 patients with a test-confirmed diagnosis of COVID-19, of whom 2,443 (51%) were women. The patient flowchart for female and male patients is depicted in **Figure 1**.

**Figure 1.**
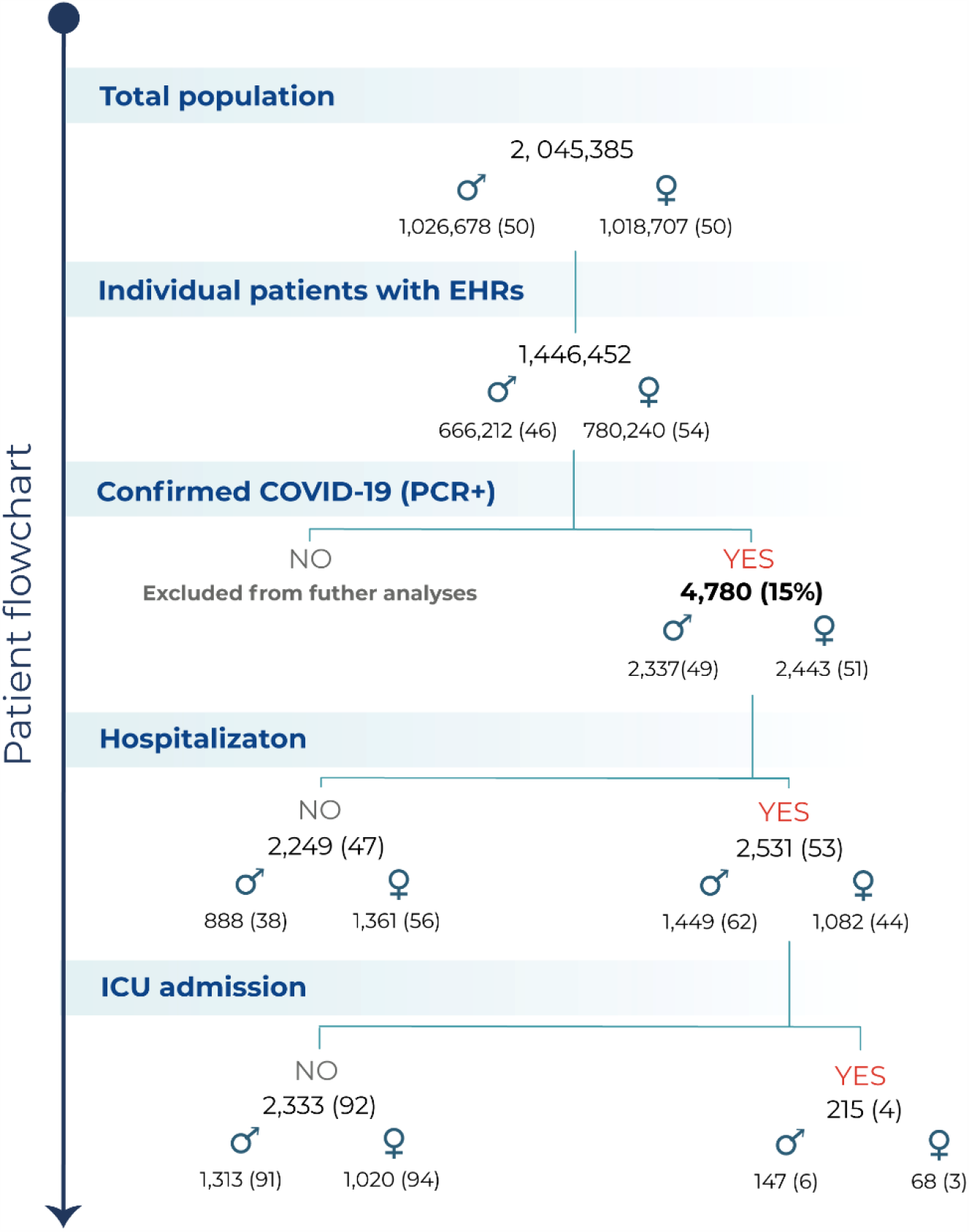
Patient flowchart. Flowchart depicting the total number of inhabitants in the source population, the number (%) of patients with available EHRs analyzed, the number of patients diagnosed with COVID-19, and of those, the number of hospitalizations and ICU admissions. ♂ = male patients; ♀ = female patients.

Isolated COVID-19 cases were already identified in the SESCAM system early in January and February 2020, yet they were scarce up to the first week of March 2020. Shortly after, confirmed cases raised exponentially and reached a daily maximum at the end of March/early April, 2020. This peak in newly reported cases was followed by a slow decrease; by early May 2020, confirmed cases went close to near-zero levels (**Figure 2a**). As shown in **Figure 2b**, the proportion of COVID-19 cases in females remained remarkably constant throughout the beginning of the outbreak up to the plateau, while it significantly increased by the end of the study period. Our linear regression analyses showed that the sex ratio of confirmed cases (newly identified cases in females over new cases in males) significantly increased over time, p < 0.001.

**Figure 2.**
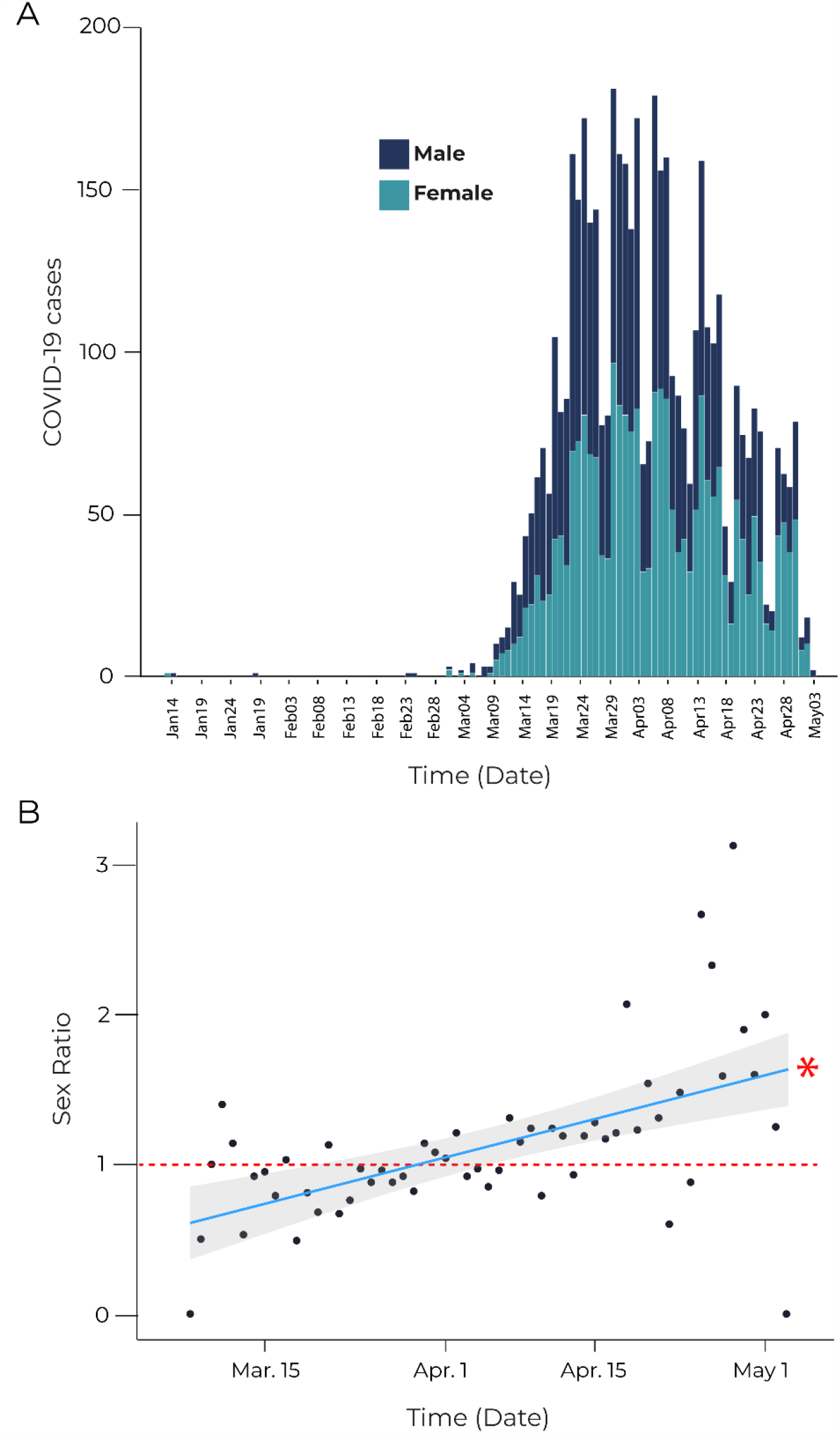
Epidemiological curve and sex ratios showing COVID-19 cases within the study period. (A) Epidemiological curve showing test-confirmed COVID-19 cases (i.e., PCR+/ IgG/IgM+) across time within the study period in male (blue) and female (green) patients. (B) Sex Ratios depicting the variation of confirmed COVID-19 cases over time within the study period, calculated as the number of diagnosed female patients over male patients. The dotted red line indicates a sex ratio of 1 (that is, equal proportion of diagnosed male and female patients). As indicated by the linear regression plot, the sex ratio increases over time, indicating a growing number of diagnosed women (in relation to men). *p < 0.001 (slope). Shaded gray area indicates CI (95%).

Female COVID-19 patients were on average 1.5 years younger than males (61.7±19.4 vs. 63.3±18.3, p=0.0025). In addition, there were more female patients in the 15-59 yr-old interval (**Figure 3**), with the greatest sex ratio (SR; 95% CI) observed in the 30-39 yr.-old interval (1.686; 1.351-2.113) **(Table 1)**.

**Figure 3.**
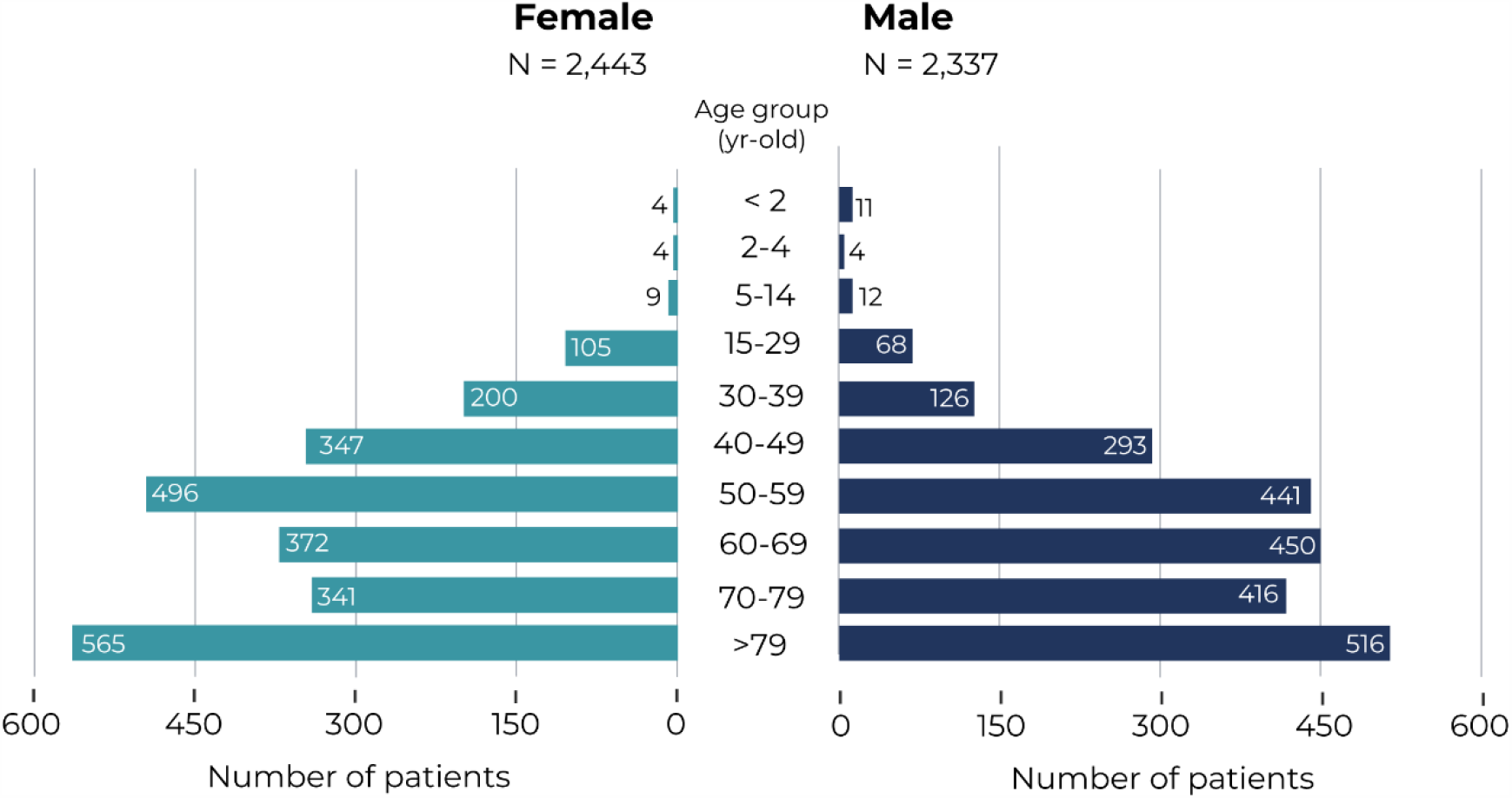
Age and Sex Distribution of COVID-19 patients. Age distribution of incident cases of COVID-19 in females (left) and males (right) in the study population for the period comprised between Jan 1, 2020 and May 1, 2020.

**TABLE 1.** Number of COVID-19 cases by age group and sex.

| Age (yr-old) | Total population* |  | Cases<br>(per 100,000) |  | Sex ratio | IC (95%) | p-value** |
| --- | --- | --- | --- | --- | --- | --- | --- |
|  | Female | Male | Female | Male |  |  |  |
| **Total** | 1,018,707 | 1,026,678 | 239.7 | 227.6 | 1.054 | 0.995-1.115 | 0.0741 |
| <15 | 148,133 | 157,505 | 11.5 | 17.1 | 0.672 | 0.358-1.225 | 0.2486 |
| 15-29 | 156,432 | 168,664 | 67.1 | 40.3 | 1.663 | 1.229-2.267 | 0.0012 |
| 30-39 | 128,166 | 136,230 | 156.0 | 92.5 | 1.686 | 1.351-2.113 | <0.001 |
| 40-49 | 159,660 | 169,961 | 217.3 | 172.4 | 1.261 | 1.079-1.474 | 0.0039 |
| 50-59 | 150,689 | 157,227 | 329.2 | 280.5 | 1.173 | 1.032-1.335 | 0.0159 |
| 60-69 | 108,557 | 109,862 | 342.7 | 409.6 | 0.837 | 0.729-0.960 | 0.0121 |
| 70-79 | 85,197 | 73,926 | 400.2 | 562.7 | 0.711 | 0.616-0.821 | <0.001 |
| >79 | 81,970 | 53,301 | 689.3 | 968.1 | 0.712 | 0.632-0.803 | <0.001 |

**Footnote:**\*Total population of Castilla La-Mancha (Spain). \*\*p-values from Yates-corrected $\chi^2$ test on percentage difference of female vs. male COVID-19 patients.

We did not observe any sex-dependent differences in the number of COVID-19 cases per 100,000 individuals; the prevalence rates for female and male patients was 239.7 and 227.6, respectively, with a corresponding sex ratio (95% CI) of 1.054 (0.995-1.115), p=0.0741 **(Table 1)**. The data shown in **Table 2** indicates an age-dependent increase in reported cases in both males and females, being patients aged >79 years the most affected with rates of 968.1 in men and 689.3 in women, and corresponding sex ratio (95% CI) of 0.712 (0.632-0.803), p<0.001.

**TABLE 2.** Clinical manifestations of COVID-19 upon diagnosis.

|  | Female, n(%)<br>(N=2,443) | Male, n(%)<br>(N=2,337) | TOTAL, n(%)<br>(N=4,780) | Sex<br>ratio | 95%CI | p-value* |
| --- | --- | --- | --- | --- | --- | --- |
| **Age (yr-old)** |  |  |  |  |  |  |
| Mean(SD) | 61.7(19.4) | 63.3 (18.3) | 62.5 (18.9) |  |  | 0.0025 |
| **Signs and Symptoms, n (%)** |  |  |  |  |  |  |
| Patients with no symptoms | 705(28.9) | 486(20.8) | 1191(24.9) | 1.387 | 1.22-1.579 | <0.001 |
| Cough | 1,094 (44.8) | 1,199 (51.3) | 2,293 (48.0) | 0.873 | 0.79-0.964 | 0.0080 |
| Fever | 878 (36.0) | 1,169 (50.0) | 2,047 (42.8) | 0.719 | 0.647-0.797 | <0.001 |
| Dyspnoea | 759 (31.1) | 914 (39.1) | 1,673 (35.0) | 0.794 | 0.71-0.888 | <0.001 |
| Respiratory crackles | 472 (19.3) | 627 (26.8) | 1,099 (23.0) | 0.720 | 0.631-0.822 | <0.001 |
| Diarrhoea | 385 (15.8) | 350 (15.0) | 735 (15.4) | 1.052 | 0.901-1.23 | 0.5467 |
| Headache | 277 (11.3) | 166 (7.1) | 443 (9.3) | 1.596 | 1.307-1.953 | <0.001 |
| Myalgia | 230 (9.4) | 207 (8.9) | 437 (9.1) | 1.063 | 0.874-1.294 | 0.5757 |
| Lymphopenia | 147 (6.0) | 186 (8.0) | 333 (7.0) | 0.756 | 0.604-0.945 | 0.0163 |
| Rhonchus | 133 (5.4) | 179 (7.7) | 312 (6.5) | 0.711 | 0.563-0.896 | 0.0044 |
| Chest pain | 158 (6.5) | 153 (6.5) | 311 (6.5) | 0.988 | 0.785-1.243 | 0.9635 |
| Anosmia | 153 (6.3) | 109 (4.7) | 262 (5.5) | 1.342 | 1.044-1.731 | 0.0254 |
| Tachypnoea | 74 (3.0) | 133 (5.7) | 207 (4.3) | 0.533 | 0.397-0.71 | <0.001 |
| Wheezing | 69 (2.8) | 86 (3.7) | 155 (3.2) | 0.768 | 0.555-1.059 | 0.1250 |
| Skin symptoms (*) | 39(1.6) | 34(1.5) | 73(1.5) | 1.097 | 0.689-1.753 | 0.7834 |
| Rhinitis | 24 (1.0) | 24 (1.0) | 48 (1.0) | 0.957 | 0.538-1.701 | 0.9938 |
| Ageusia | 31(1.3) | 15 (0.6) | 46 (1.0) | 1.966 | 1.073-3.766 | 0.0403 |
| Sore throat | 27 (1.1) | 18 (0.8) | 45 (0.9) | 1.431 | 0.789-2.657 | 0.2993 |
| Dysphagia | 12 (0.5) | 20 (0.9) | 32 (0.7) | 0.577 | 0.272-1.173 | 0.1746 |
| Neuralgia | 16 (0.7) | 13 (0.6) | 29 (0.6) | 1.175 | 0.561-2.507 | 0.8025 |
| Hemoptysis | 9 (0.4) | 12 (0.5) | 21 (0.4) | 0.721 | 0.29-1.723 | 0.5919 |
| Ophthalmologic symptoms (#) | 9(0.4) | 9(0.4) | 18(0.4) | 0.957 | 0.368-2.487 | 1 |
| Splenomegaly | 3 (0.1) | 6 (0.3) | 9 (0.2) | 0.491 | 0.098-1.924 | 0.4641 |
| Hepatomegaly | 2 (0.1) | 5 (0.2) | 7 (0.1) | 0.400 | 0.051-1.943 | 0.4159 |
| **Respiratory rate (bpm)** |  |  |  |  |  |  |
| N | 249 | 339 | 588 |  |  |  |
| Mean(SD) | 23.3(14.4) | 23.9(12.6) | 23.7(13.4) |  |  |  |
| Patients (n,%) with high RR (>20) | 105(42.3) | 167(49.3) | 272(46.3) | 0.856 | 0.637-1.148 | 0.3356 |
| **Radiological findings, n (%)** |  |  |  |  |  |  |
| Chest X-ray | 1600(65.5) | 1829(78.3) | 3429(71.7) | 0.837 | 0.766-0.914 | <0.001 |
| No abnormalities | 552(34.5) | 450(24.6) | 1002(29.2) | 1.402 | 1.217-1.615 | <0.001 |
| Any abnormality | 1048(65.5) | 1379(75.4) | 2427(70.8) | 0.869 | 0.782-0.965 | 0.0091 |
| Bilateral infiltrates | 902(56.4) | 1213(66.3) | 2115(61.7) | 0.850 | 0.762-0.948 | 0.0039 |
| Ground-glass opacities | 277(17.3) | 380(20.8) | 657(19.2) | 0.833 | 0.704-0.986 | 0.0378 |
| Interstitial pattern | 152(9.5) | 175(9.6) | 327(9.5) | 0.993 | 0.790-1.246 | 0.9972 |
| Alveolar bilateral infiltrates | 60(3.8) | 88(4.8) | 148(4.3) | 0.780 | 0.556-1.088 | 0.1683 |
| Unilateral infiltrates | 7(0.4) | 15(0.8) | 22(0.6) | 0.540 | 0.203-1.295 | 0.2393 |
| **Arterial blood gases, n (%)** |  |  |  |  |  |  |
| **pH** |  |  |  |  |  |  |
| N | 476 | 646 | 1122 |  |  |  |
| Mean(SD) | 7.4(0.1) | 7.4(0.1) | 7.4(0.1) |  |  |  |
| Patients (n,%) with pH>7.42 | 298(62.6) | 449(69.5) | 747(66.6) | 0.901 | 0.746-1.087 | 0.2982 |
| **pO<sub>2</sub> (mmHg)** |  |  |  |  |  |  |
| N | 553 | 778 | 1331 |  |  |  |
| Mean(SD) | 72.1(24.8) | 70.5(26.9) | 71.2(26.1) |  |  |  |
| Patients (n,%) with pO<sub>2</sub><60 | 144(26.0) | 249(32.0) | 393(29.5) | 0.814 | 0.644-1.026 | 0.0924 |
| **pCO<sub>2</sub> (mmHg)** |  |  |  |  |  |  |
| N | 443 | 622 | 1065 |  |  |  |
| Mean(SD) | 35.8(7.3) | 34.5(7.9) | 35.1(7.7) |  |  |  |
| Patients (n,%) with pCO<sub>2</sub>>45 | 46(10.5) | 42(6.8) | 88(8.4) | 1.538 | 0.993-2.387 | 0.0661 |
| **O<sub>2</sub> Sat (%)** |  |  |  |  |  |  |
| N | 1188 | 1336 | 2524 |  |  |  |
| Mean(SD) | 94.1(5.7) | 93.3(5.6) | 93.7(5.6) |  |  |  |
| Patients (n,%) with O<sub>2</sub> Sat<94 | 385(32.4) | 528(39.5) | 913(36.2) | 0.820 | 0.704-0.955 | 0.0122 |

**Footnote:** \*p-values from Yates-corrected chi<sup>2</sup> test of difference between percentage of patients (female vs. male) presenting with the sign/symptom. All tests were performed individually for each variable sign/symptom.

Regarding symptoms upon diagnosis, headache, anosmia, and ageusia were significantly more frequent in women than men, all p<0.001 (**Table 2**). Interestingly, imaging by chest X-ray or blood tests were performed less frequently in females (65.5% vs. 78.3% and 49.5% vs. 63.7%, respectively), all p<0.001. Regarding hospital resource use, female COVID-19 patients showed less frequency of hospitalization (44.3% vs. 62.0%) and ICU admission (2.8% vs. 6.3%) than males, all p<0.001.

As expected, comorbidities upon COVID-19 diagnosis were more often reported in men. Whereas 78.9% of female patients had at least one of the studied comorbidities at diagnosis, this percentage was 87.4% in males (p=0.0183) (**Table 3**). However, depressive disorders and asthma were significantly more frequent in females, with associated ratios of 2.030 (1.616-2.565) and of 1.743 (1.363-2.241), respectively.

**TABLE 3.**
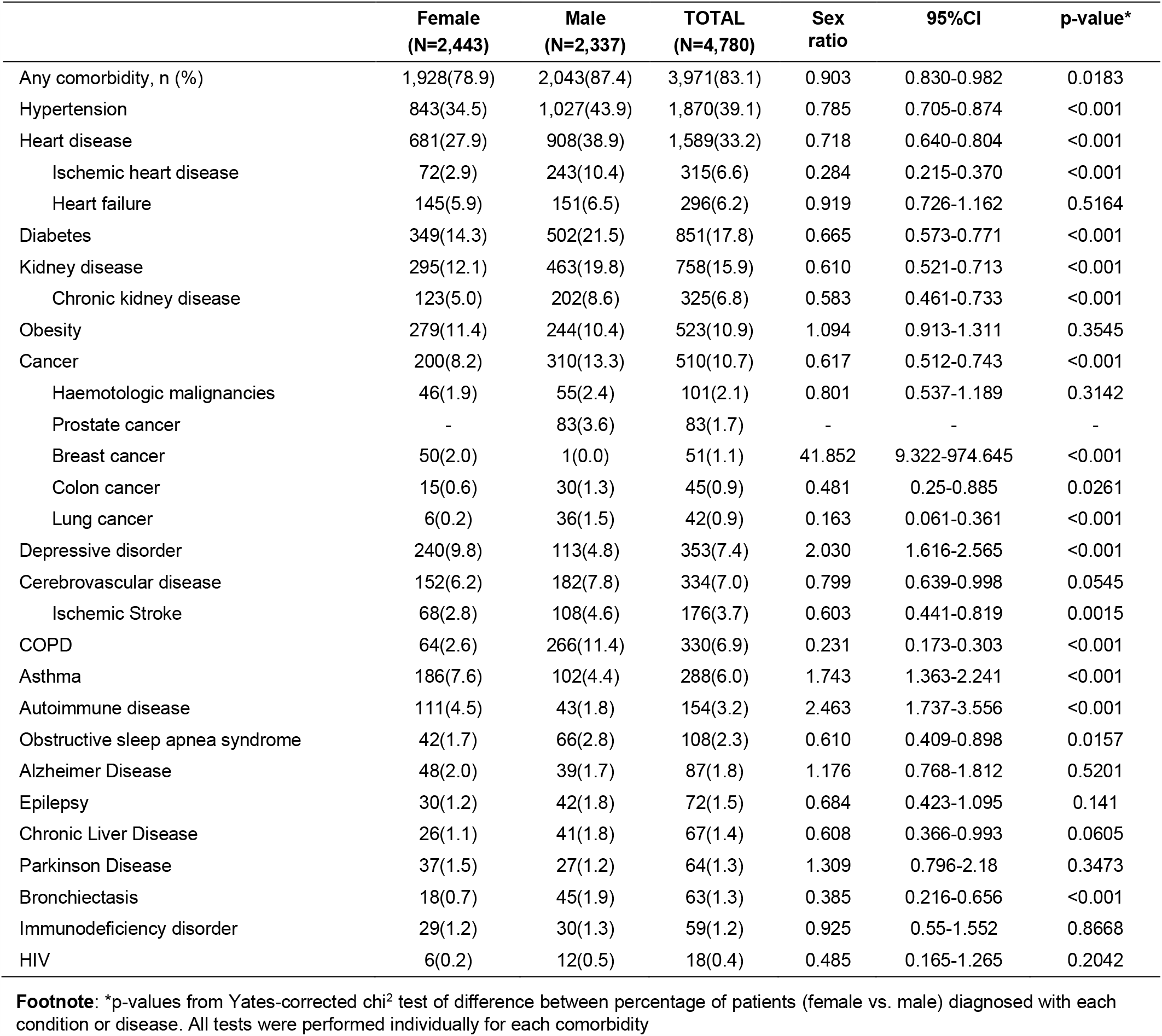
Comorbidities of COVID-19 patients upon diagnosis.

According to the laboratory parameters upon COVID-19 diagnosis, men significantly suffered more from lymphopenia and worse renal function (as per creatinine and urea values but not GFR) than women (**Table 4**). Au contraire, all liver function parameters, as well as D-dimer and all acute phase reactants (except for higher CRP levels in men) were also evenly distributed by sex.

**TABLE 4.**
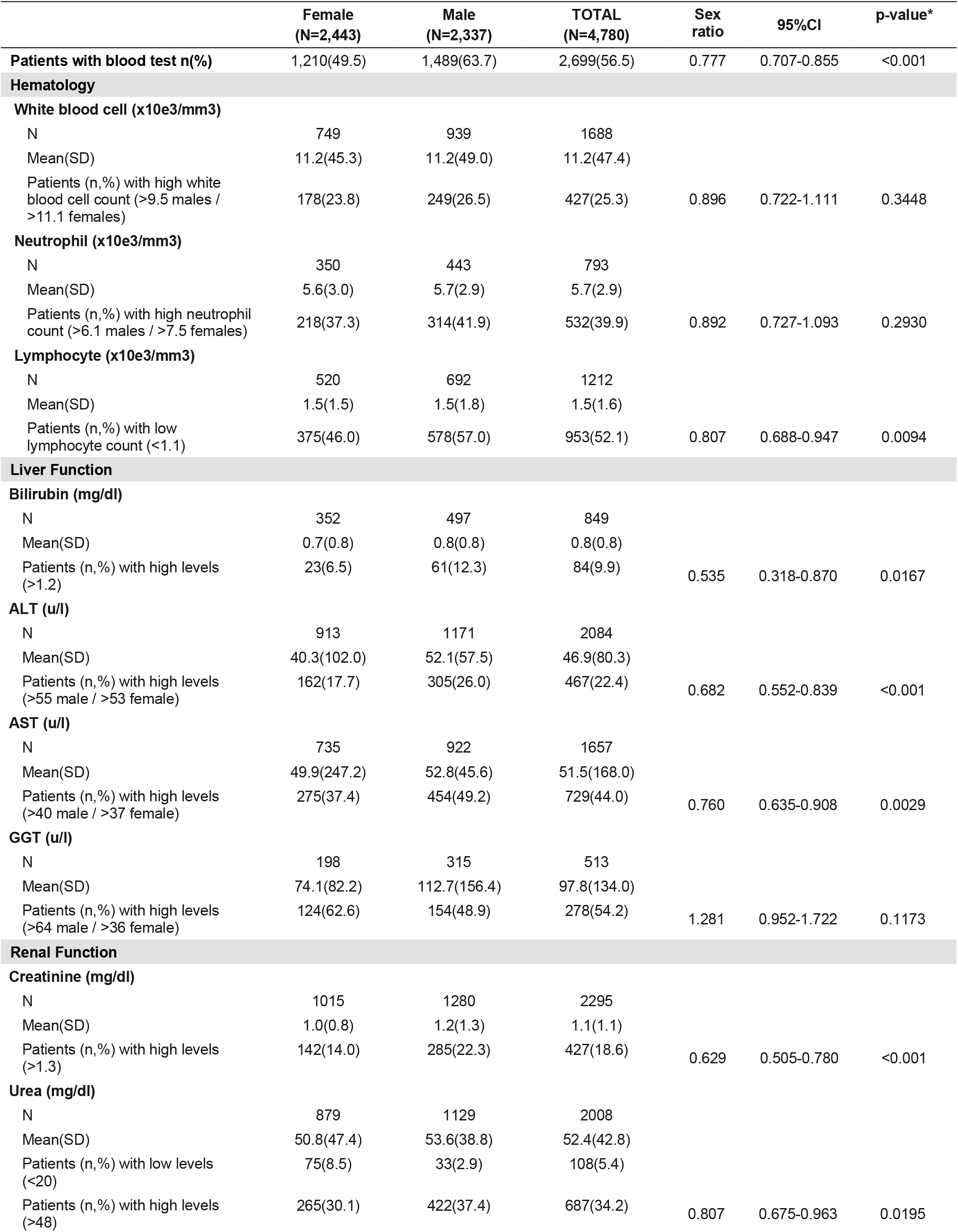
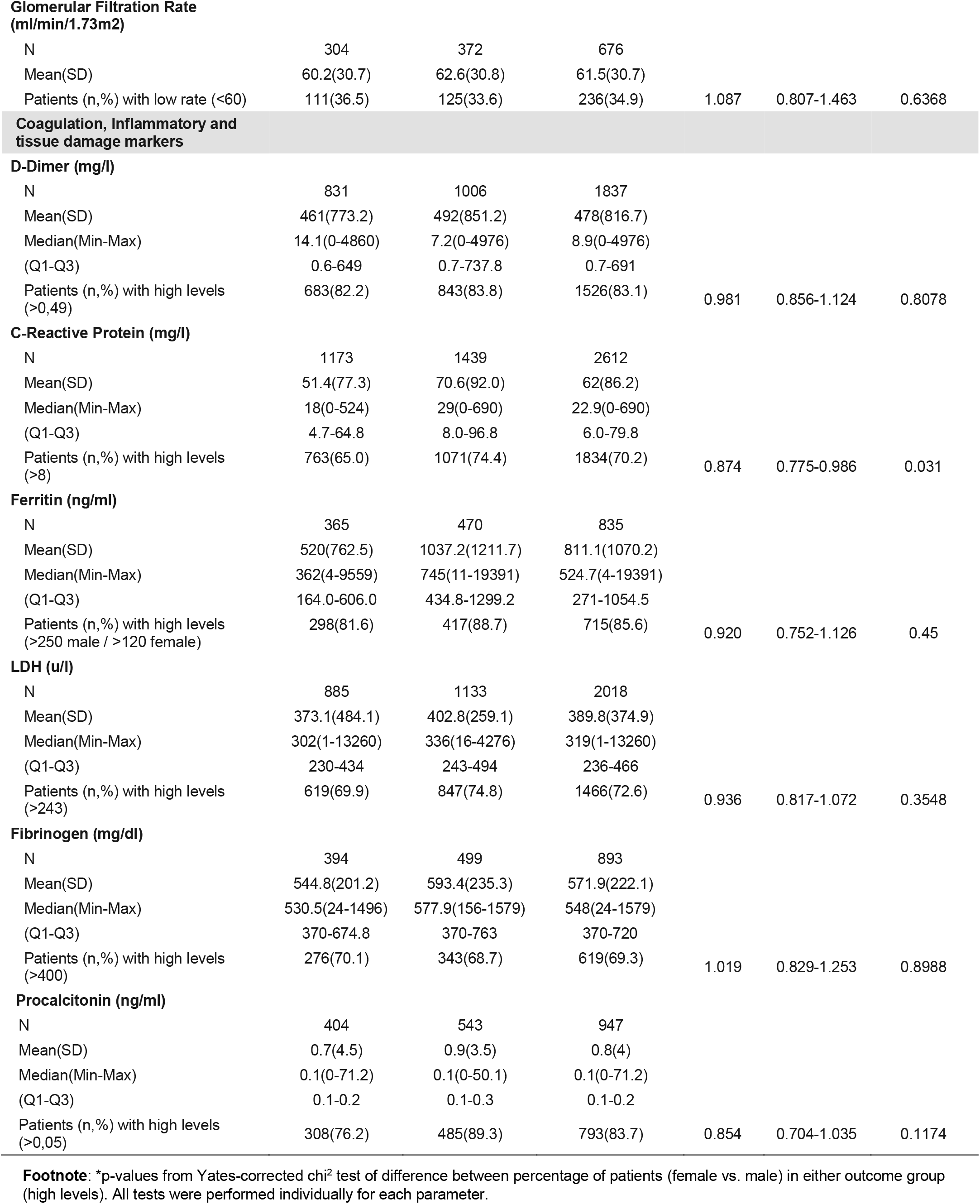
Laboratory parameters of COVID-19 patients upon diagnosis.

Finally, we explored the treatments received by all COVID-19 patients (**Table 5**). Our results indicate that except chloroquine, the sex ratio for all treatments analyzed was < 1. Notably, most of these comparisons were statistically significant against female patients with COVID-19 (**Table 5**).

**TABLE 5.**
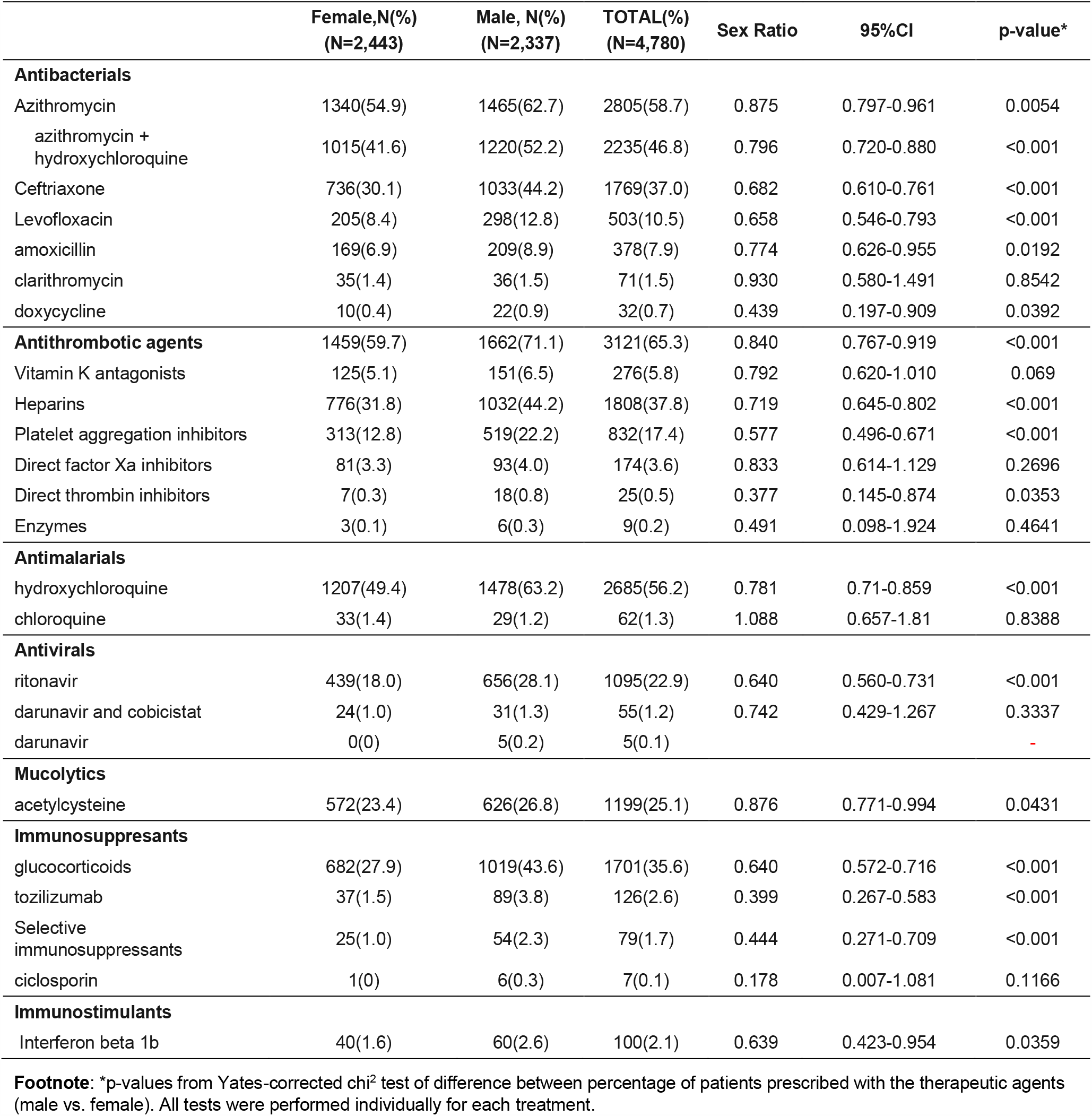
Treatments used in Covid-19 patients.

## DISCUSSION

Using a big data approach and adopting a population perspective, we have identified important sex-dependent differences in the clinical manifestation, diagnosis, management, and hospital resource use associated with COVID-19. Specifically, female teenagers and young adult women were significantly more affected by COVID-19 than their male counterparts in the same age ranges; In addition, our results indicate that headache, as well as ear, nose and throat (ENT) symptoms were significantly more frequent in female COVID-19 patients. Regarding medical outcomes, both hospitalization and ICU admission were less frequent outcomes in females than males. Unfortunately, basic diagnostic tests such as blood tests or imaging were less used in women.

Our results provide further evidence of the inherent gender bias in the Health System, which is thought to originate in medical school and impacts all aspects of healthcare. ^18 19^ Although this bias is well established the context of cardiovascular^20^, respiratory^21,22,23^, and infectious diseases (particularly, STDs^24^), the impact of sex and gender in the ongoing COVID-19 pandemic is just beginning to be unraveled.^25,26,27^ Beyond mechanistic and molecular studies,^5-9^ more subtle and general events may already play a role in the sex-dependent management of COVID-19 patients.^28,29^ One key question is whether COVID-19 affects women’s reproductive health; in other coronavirus-related infectious diseases such as the severe acute respiratory syndrome (SARS) and the middle east respiratory syndrome (MERS), pregnancy has been identified as a risk factor for developing severe complications.^30,31^

The increased vulnerability of women to COVID-19 is also associated with occupational risks. It is well established that most frontline health care professionals are women, which puts them at a higher risk for infection and negative clinical outcomes.^32^ Further, women are more likely to serve as the primary caregivers within a household, thus becoming more exposed to the disease. This becomes worrying in disadvantaged populations and resource-poor communities, as well as countries without the benefits of a universal, free-for-all healthcare system.

### Strengths and limitations

The main strengths of our research include immediacy, large sample size, and direct access to real-world evidence (RWE). Of note, our methodology ensures absence of any bias in patient selection as our hypothesis that gender impacts diagnosis and management of COVID-19 was assessed *a posteriori*. The observed change in the sex ratio of confirmed cases at the tail of this first wave of the pandemic should be further confirmed in other cohorts and geographical locations.^33^ Finally, it is unlikely that our conclusions are impacted by the limitations of pay- or copay-systems, as Spain enjoys an universal, free-for-all health care system.

Our results should be interpreted in light of the following limitations. First, given the variation in COVID-19 severity, it is possible that the free-text information available in EHRs is not homogeneous across patients seen in different points of care (i.e., primary-to-tertiary care). For instance, care providers could have been more likely to further explore (and report more often) milder symptoms in women, who in turn are more likely to be seen in primary care; on the other hand, the more sever ymptoms reported in men may be related to the fact that they were more likely to be hospitalized or visit the ICU. Second, it is possible that women were more likely than men to report ENT symptoms.^34^ Third, as indicated in the methods section, our reported COVID-19 prevalence rates are probably lower than real, as some cases might be missed by the system due to heterogeneous reporting in EHRs. However, the observed low recall metrics in variables related to the identification of PCR-confirmed patients do not affect the quality of the descriptive results since our precision metrics for these concepts were optimal.

### Implications for future research

The well-established gender bias in cardiovascular^35^, respiratory^36,37,38^, and other diseases should be further investigated in COVID-19 patients. Despite recent regulations and partial improvements, the attention paid to sex and gender differences in biomedical and health research is far from optimal.^39^ As pointed out in recent reviews, occupational gender segregation makes women particularly vulnerable to COVID-19 since two-thirds of the health and social care workforce worldwide are women.^40^ Crucially, any gender bias in the use of diagnostic testing and imaging, as evidenced in our research from a country with universal, free-for-all healthcare, might be magnified in less privileged settings.

## Conclusion

The biological, behavioral, social, and systemic factors underlying the differences in how women and men may experience COVID-19 and its consequences cannot be oversimplified.^41^ Regrettably, most research studies are systematically failing to offer comparisons between women and men, girls and boys, and people with diverse gender identities.^42^ Based on the results presented here, we conclude that women were more heavily impacted by COVID-19 than men (specifically teenagers and young adults). In addition, women presented different symptoms at disease onset, clinical outcomes, and treatment patterns. These results warrant further research to identify and close the gender gap in the diagnosis and treatment of COVID-19.

## Data Availability

Data not available due to legal restrictions

## Acknowledgments

We thank all the Savaners for helping accelerate health science with their daily work. We also thank SESCAM (Healthcare Network in Castilla-La Mancha) for its participation in the study and for supporting the development of cutting-edge technology in real time.

## Author Contribution statement for each author

JA, IHM, JLI and JBS had the original idea of the study and developed the concept protocol; AP, SL, CDRB, SM, IS and IZ developed the analytical plan and conducted the statistical analyses; CDRB and JBS wrote and edited the manuscript; CDRB and IZ are responsible for figures and data visualization; all authors contributed to drafting and interpretation, and they approved the final version.

## Author Disclosure Statement

The Big COVIData study was funded by Savana. Savana employees contributed to the design, data analysis, and writing of the present study. All authors declare there are no other direct or indirect potential conflicts to disclose.

## Notes

### Author Declarations

This study is part of the BigCOVIData initiative and was conducted in compliance with legal and regulatory requirements. This study was classified as a non-post-authorization study (EPA) by the Spanish Agency of Medicines and Health Products (AEMPS), and was approved by the Research Ethics Committee at the University Hospital of Guadalajara (Spain).

